# Low-Resource Automatic ICD Coding for Chinese Electronic Medical Records: An Interpretable Multi-View Linear Framework with Audited Clinical Rules

**DOI:** 10.64898/2026.10.04.26364688

**Authors:** Yupin Xu, Jiatao Yan

## Abstract

ICD coding is the backbone of medical-record structuring, insurance payment, and clinical research. Today’s best automatic coders are deep models, but they assume tens of thousands of labeled records, GPU servers, and machine-learning engineers. Many hospitals, especially small ones, have none of these. We study Chinese electronic medical records (EMRs) under a hard budget: 2,200 labeled records, CPU only, and fully interpretable decisions. Our method is lightweight and follows the way human coders work. First, codebook knowledge injection marks standard disease and operation names in the text as useful clues. Second, multi-view field weighting gives more weight to the clinical sections that matter most. Third, character n-gram TF-IDF features feed linear classifiers that score each candidate code. Fourth, a small set of clinical rules acts as a final safety check. Our main methodological finding is about this last step: naively applied rules do more harm than good (overall F1 drops from 0.8070 to 0.7995, and main-operation accuracy from 0.9727 to 0.9432). We therefore audit each rule on held-out data by counting how often it fixes a wrong prediction or breaks a correct one, and we remove rules that do more harm than good. This turns rule post-processing into a safety check that never hurts. On the CCL2026-Eval Task 6 dataset, our method reaches macro-F1 of 0.899 (main diagnosis) and 0.966 (main operation) in 5-fold cross-validation. A frequency-stratified analysis shows a clear long-tail limit: codes seen fewer than three times score zero F1. In a head-to-head test, a fine-tuned Chinese pretrained encoder scores lower than our method on all four tasks at this training size, while needing a GPU. Ablation tests show that the gains come mainly from field-weighted text. The framework trains in about 40 minutes on an ordinary CPU, and every prediction can be broken down into interpretable feature contributions.

## 1. Problem and Motivation

ICD (International Classification of Diseases) coding gives each clinical record standardized diagnosis and procedure codes. It is the base of hospital information systems, insurance payment, epidemiology, and clinical research. In most hospitals, and in nearly all small community hospitals, coding is still done by hand: trained coders read each record and pick codes from national codebooks. This work is expensive, slow, and different coders often disagree.

Ten years of research have produced strong deep-learning coders, from convolutional label-attention models [1, 2] to pretrained-language-model coders [3]. These results rely on three things that most of the world does not have: large labeled datasets (MIMIC-III holds over 50,000 English discharge summaries [4]), GPU servers, and engineers who can run deep models. Recent replication studies also show that many reported gains are fragile [5]. For a small hospital with a few thousand old records, no GPU, and no machine-learning staff, this literature is hard to use in practice. Such hospitals are not rare; they are the norm in low- and middle-income regions.

This work began as the two authors’ joint entry to the CCL2026-Eval Task 6 competition, which ranked 13th among 225 teams. The competition made the low-resource constraints concrete, and it convinced us that these research questions deserve deeper study.

So this paper asks a practical question: *can we close this gap by improving the algorithm, instead of collecting more data or buying more compute?* More precisely: *with only 2,200 labeled Chinese EMRs, a CPU, and no deep-learning experts, how far can a carefully built classical pipeline go, and can every step be kept clear and trustworthy?* Our answer is a qualified yes, with clear limits. Head codes can be coded automatically with a few thousand records and ordinary hardware. Rare codes stay out of reach for information-theoretic reasons. Expert rules are safe only after each rule is checked on real data. Notably, at this data size a fine-tuned Chinese pretrained encoder does not even match our lightweight pipeline. This tells us that the low-resource gap is at least partly an algorithm problem, not only a data problem.

## 2. Study Overview

Figure 1 sums up the whole study in four panels: the data and tasks (a), the five-step pipeline (b), the evaluation protocols and comparison methods (c), and the main measured results (d). Panel a: 2,200 Chinese EMRs (1,800 from the official training release plus 400 evaluation records with released labels), 15 text fields per record, four coding targets, and a long tail. Panel b: a CPU-only pipeline that follows the way human coders work: codebook knowledge injection (1), multi-view field-weighted text construction (2), character n-gram TF-IDF with linear classifiers (3), an audited clinical-rule safety check (4), and the four-code output (5). Panel c: two evaluation protocols (8:2 hold-out and 5-fold cross-validation), plus learning-curve, ablation, efficiency, and interpretability analyses, and the comparison methods (classical baselines and a fine-tuned deep encoder). Panel d: the main results: usable head-code accuracy (0.925/0.973), rules that go from harmful to safe after auditing, and a fine-tuned encoder that scores lower than our pipeline on all four tasks at this size.

**Figure 1.**
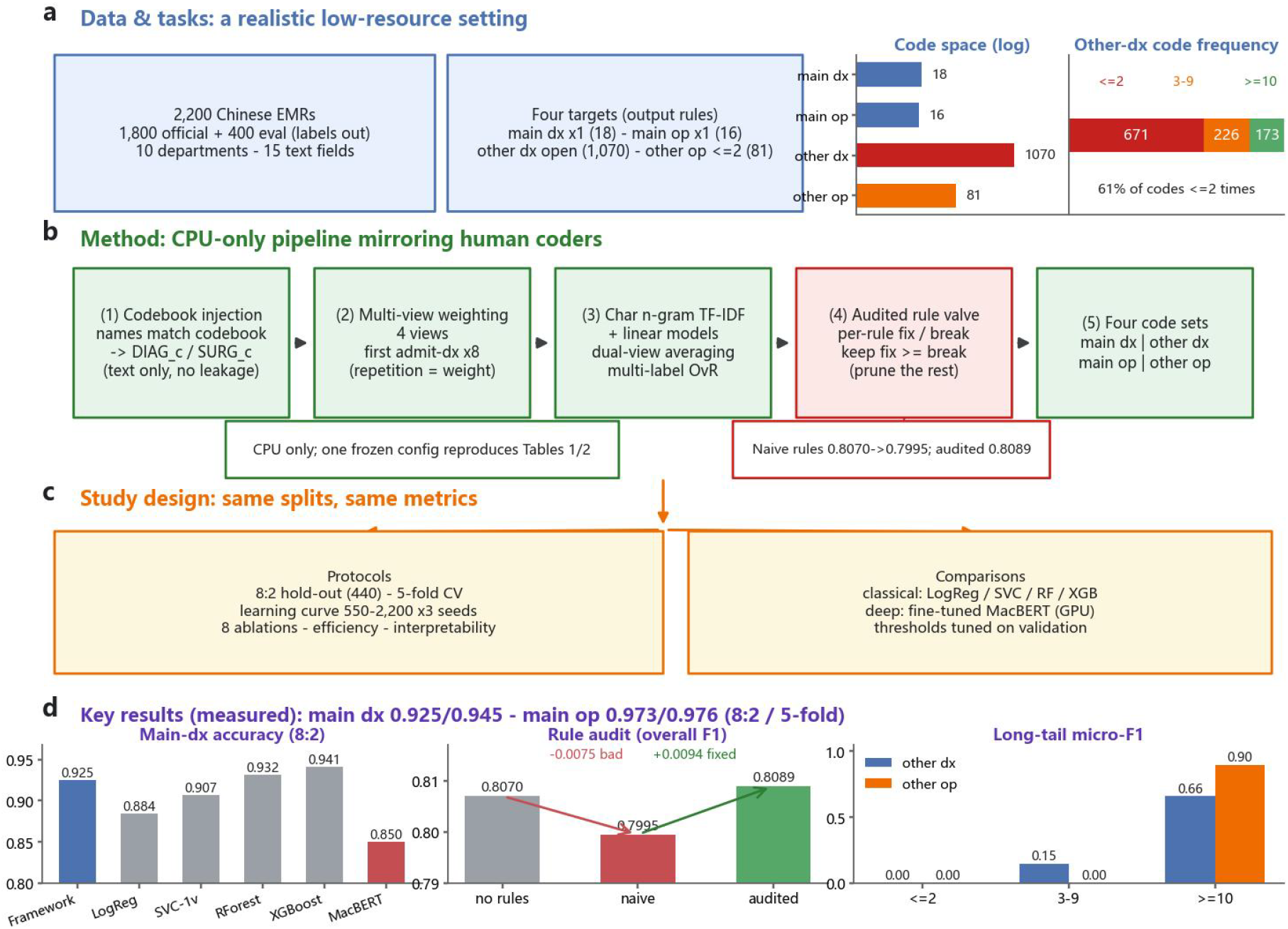
Study overview. (a) Data and tasks of the low-resource setting. (b) The CPU-only five-step pipeline, including the audited rule safety check. (c) Evaluation protocols and comparison methods. (d) Main measured results. Every number in the figure comes from the experiments in Section 4.

Three design rules guide the framework. *Data-efficient*: every part can be trained on the few thousand records a hospital already owns. No external corpus, no pretrained weights, no GPU. *Interpretable*: every decision is a fixed, checkable transformation. The final score of a code adds up from character n-gram contributions that we can list. *Auditable*: expert knowledge enters only through channels we can check. The codebook channel never uses labels, and every clinical rule carries a measured fix/break record before it is used. We report one frozen hyperparameter setting throughout; this makes deployment, auditing, and reproduction simpler.

## 3. Method and Data

### 3.1 Data source and structure

We use the CCL2026-Eval Task 6 dataset (Chinese EMR ICD coding evaluation; Tianchi competition #532466). It was built by the QLU-NLP group (Qilu University of Technology) and the First Affiliated Hospital of Shandong First Medical University from de-identified records [9]. We use the 2,200 labeled records we can access (1,800 from the official training release plus 400 evaluation records with released labels). Every number in this paper comes from reproducible local runs on this set. The data cover 10 clinical departments. Each record has 15 text fields, including chief complaint, history of present illness, admission diagnosis, operative notes, and treatment course. Four targets are annotated: *main diagnosis* (single-label; 18 unique codes; ICD-10 NCE 2.0), *main operation* (single-label; 16 unique codes; ICD-9-CM3 NCE 3.0), *other diagnoses* (multi-label; 1,070 unique codes in training), and *other operations* (multi-label; 81 unique codes). The label distribution has a long tail: about 61% of other-diagnosis codes appear fewer than three times in training. This tail, not the head codes, drives the macro-averaged metrics; it is the main difficulty of the multi-label tasks (Section 4.5). The evaluation format requires exactly one main diagnosis and one main operation per record, at most two other-operation codes, and a “;”-separated list of other-diagnosis codes with no fixed length. Gold sets contain at most 8 and 2 codes, so our top-k cut-offs (k=10,2) never remove any gold label. We score an empty prediction against an empty gold set as F1 = 1, and we state this rule wherever sample-F1 appears.

### 3.2 Processing steps (following the roadmap)

1. **Codebook knowledge injection**. Human coders read the codebook; we let the model do the same. From the ICD-10 / ICD-9-CM3 codebooks and our alias tables, we build a *code → names* index for every code seen in training (the standard name plus known aliases). If any name of code c appears in a record’s diagnosis or operation fields, we add a token `DIAG_c` or `SURG_c` to the text. The procedure uses only input text, never labels, so it works at a new hospital without leakage. (Section 4 reports a null result for this channel on our dataset; we keep it as a clean knowledge path.)
2. **Multi-view field-weighted text construction**. Fields differ in how much they tell us about each task. We group the 15 fields into diagnosis-key fields (admission, pre-operative, intra-operative, discharge diagnoses), operation-key fields (operative notes, treatment course, pre-/intra-operative diagnoses), and the full set. Then we build four *views* by repeating field groups a task-specific number of times. Under TF-IDF, repetition is how we set field weights. For example, the diagnosis-focused view repeats the first admission-diagnosis entry eight times, because it is the strongest evidence for the main diagnosis.
3. **Character n-gram TF-IDF**. Each view is turned into numbers with character n-gram TF-IDF (n ∈ [2,4], sublinear TF). Character features need no word segmenter, and they handle the wording differences of Chinese clinical text: different spellings of the same term (short forms versus full names) still share character substrings.
4. **Linear decision models**. For the two single-label tasks, we train two `LinearSVC` classifiers (`class_weight=balanced`) on two views. The final score is the average of the two decision functions. A small pairwise discriminator (an override with a threshold) handles codes that are easy to confuse. The multi-label tasks use `OneVsRest(LinearSVC)` over codes with training frequency ≥ 2 / ≥ 3. Prediction keeps codes above a decision threshold and takes the top-k by score. Two statistics-based layers then raise recall on the long tail: conditional-frequency supplementation (adds codes with P(code | main diagnosis) ≥ 0.65, estimated on training data) and term-to-code mapping (maps standard terms from diagnosis fields to codes above frequency and probability gates). Frozen hyperparameters: C = 3.6/2.4 (main-diagnosis views, 260k/220k features), C = 1.2/3.6 (main operation, 180k/260k), C=10 with thresholds -0.40 / -0.20 and k=10 / 2 for the multi-label tasks, minimum frequencies 2 / 3.
5. **Auditable rule-based verification**. A final layer of hand-written clinical rules (for example, chemotherapy context → Z51.x codes with 99.2503; prostatic laser resection → N40.x00) adds clinical constraints to model outputs. Our methodological point: such rules *must be checked before use*. For each rule r, we collect the records whose prediction it overrides on a held-out split, and we count

*fix_r = #{override changes wrong → right}, break_r = #{override changes right → wrong}*.

We keep rule r only if its net effect Δ_r = fix_r - break_r ≥ 0. Rules with negative net effect are removed or restricted. We state two caveats. First, auditing many rules on one split risks overfitting that split; nested or repeated splits are the right fix. Second, when a rule fires rarely, Δ_r is noisy. So borderline rules (Δ_r = 0) are kept only if they enforce code-space legality (removing duplicated or cut-off codes), which carries no clinical risk.

## 4. Results

### 4.1 Main results and the rule audit

Tables 1 and 2 agree with each other. The same frozen setting under two protocols gives main-diagnosis accuracy of 0.925 (hold-out) / 0.945 (5-fold), and main-operation accuracy of 0.973 / 0.976. The 5-fold rule ablation confirms that the v2 audit is safe: main-operation accuracy is the same with and without rules (0.9764); main-diagnosis accuracy rises a little with rules (0.9445 vs.\ 0.9405); multi-label F1 does not change (0.5690 / 0.8836). The pipeline contains no random components: with the frozen configuration, repeated runs reproduce every number in Tables 1–3 exactly.

**Table 1.** Held-out results (8:2, seed 42). Acc = accuracy; F1 = macro-F1 for single-label tasks and sample-F1 for multi-label tasks. Naive rules do more harm than good; audited rules (v2) remove that harm and slightly raise main-diagnosis F1 over the rule-free base.

| Method | Main-Diag |  | Main-Op |  | O-Diag | O-Op | Overall |
| --- | --- | --- | --- | --- | --- | --- | --- |
|  | Acc | F1 | Acc | F1 | F1 | F1 | (mean F1) |
| Base (no rules) | 0.9182 | 0.8425 | 0.9727 | 0.9592 | 0.5573 | 0.8689 | 0.8070 |
| + naive rules (v1) | 0.9136 | 0.8407 | 0.9432 | 0.9312 | 0.5569 | 0.8689 | 0.7995 |
| **+ audited rules (v2)** | **0.9250** | **0.8502** | **0.9727** | **0.9592** | **0.5573** | **0.8689** | **0.8089** |

**Table 2.** 5-fold cross-validation of the final framework (audited rules, v2), averaged over 5 folds. Single-label tasks report macro-P/R/F1; multi-label tasks report sample-level P/R/F1.

| Task | Acc | P | R | F1 |
| --- | --- | --- | --- | --- |
| Main-Diag | 0.9445 | 0.9025 | 0.8967 | 0.8989 |
| Main-Op | 0.9764 | 0.9676 | 0.9662 | 0.9662 |
| O-Diag | — | 0.6432 | 0.5556 | 0.5690 |
| O-Op | — | 0.8939 | 0.8820 | 0.8836 |
| Composite (mean F1) |  |  |  | **0.8294** |

The audit record (Table 3) is the main methodological evidence. The biggest single harm came from the gastric-polyp/gastroscopy rule family: it overwrote 14 correct `43.4105` (polypectomy) predictions with `45.1300×004` (gastroscopy), while fixing only 2. Two auto-completion rules were harmful as well. After pruning, the rules that remain are code-space cleanups and corrections with no negative effect.

**Table 3.**
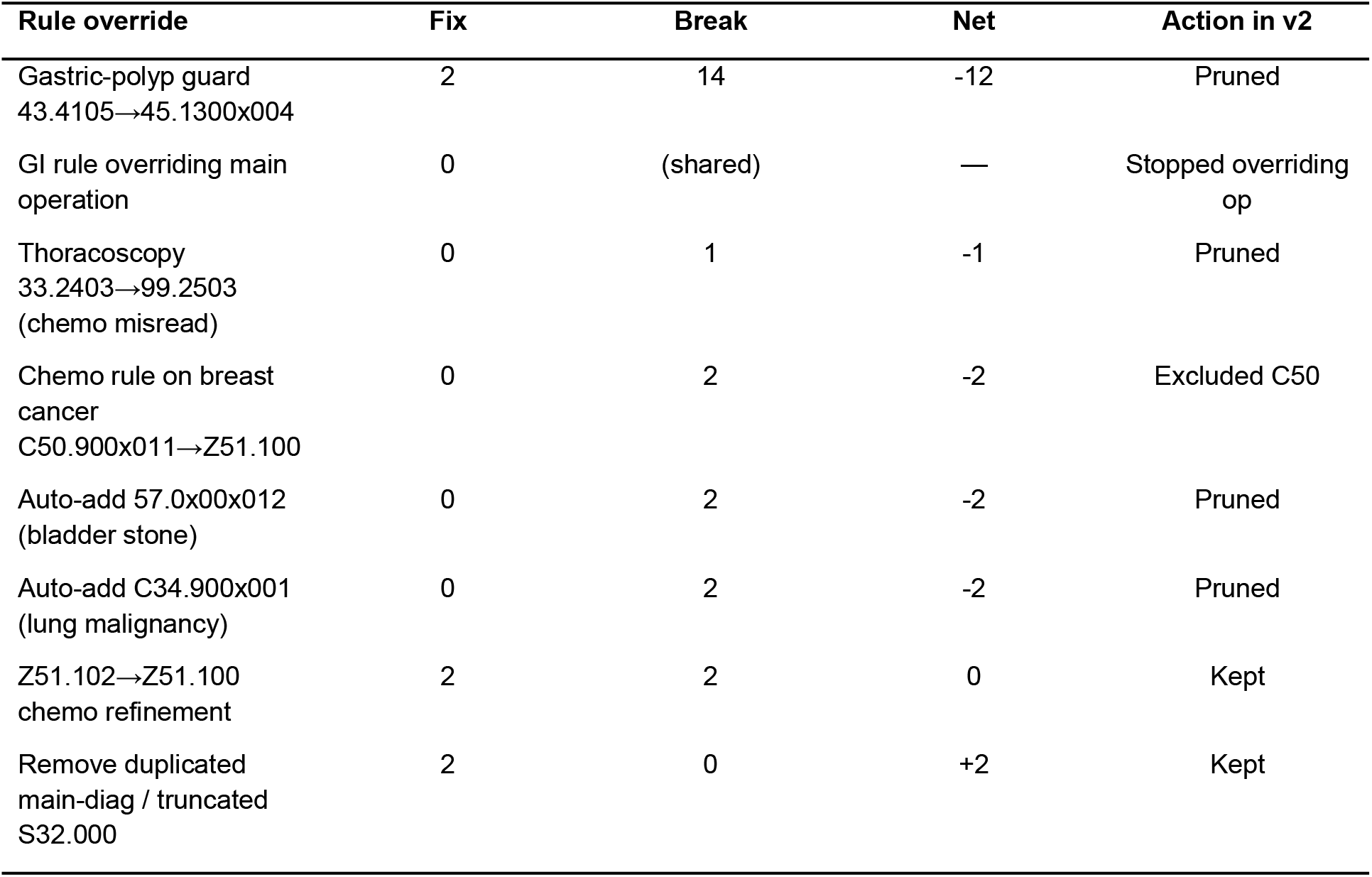
Per-rule audit on the validation split (fix = wrong→correct overrides; break = correct→wrong). Rules with negative net effect are removed in v2.

### 4.2 Module ablations

Table 4 reports one-factor ablations on the held-out split. The dominant factor is *field weighting*: removing it costs 2.0 points of overall F1, including 5.3 points of other-diagnosis F1 and 0.9 points of main-operation accuracy. Multi-view fusion adds a modest 0.2 points. Three parts bring no measurable gain. Codebook knowledge injection leaves both single-label tasks bit-identical. Replacing character n-grams with jieba word features is a tie (0.8083 vs.\ 0.8089). The pairwise discriminator is neutral to slightly negative (removing it changes overall F1 by +0.13 points, which is noise on 440 records). Removing the conditional-frequency supplement is neutral as well (+0.03). The audited rule layer has no negative effect (+0.19 overall, driven by +0.68 main-diagnosis accuracy), as its design intends. Differences below one point on 440 records should be read with care; the field-weighting effect is ten times larger and is the framework’s main lever.

**Table 4.**
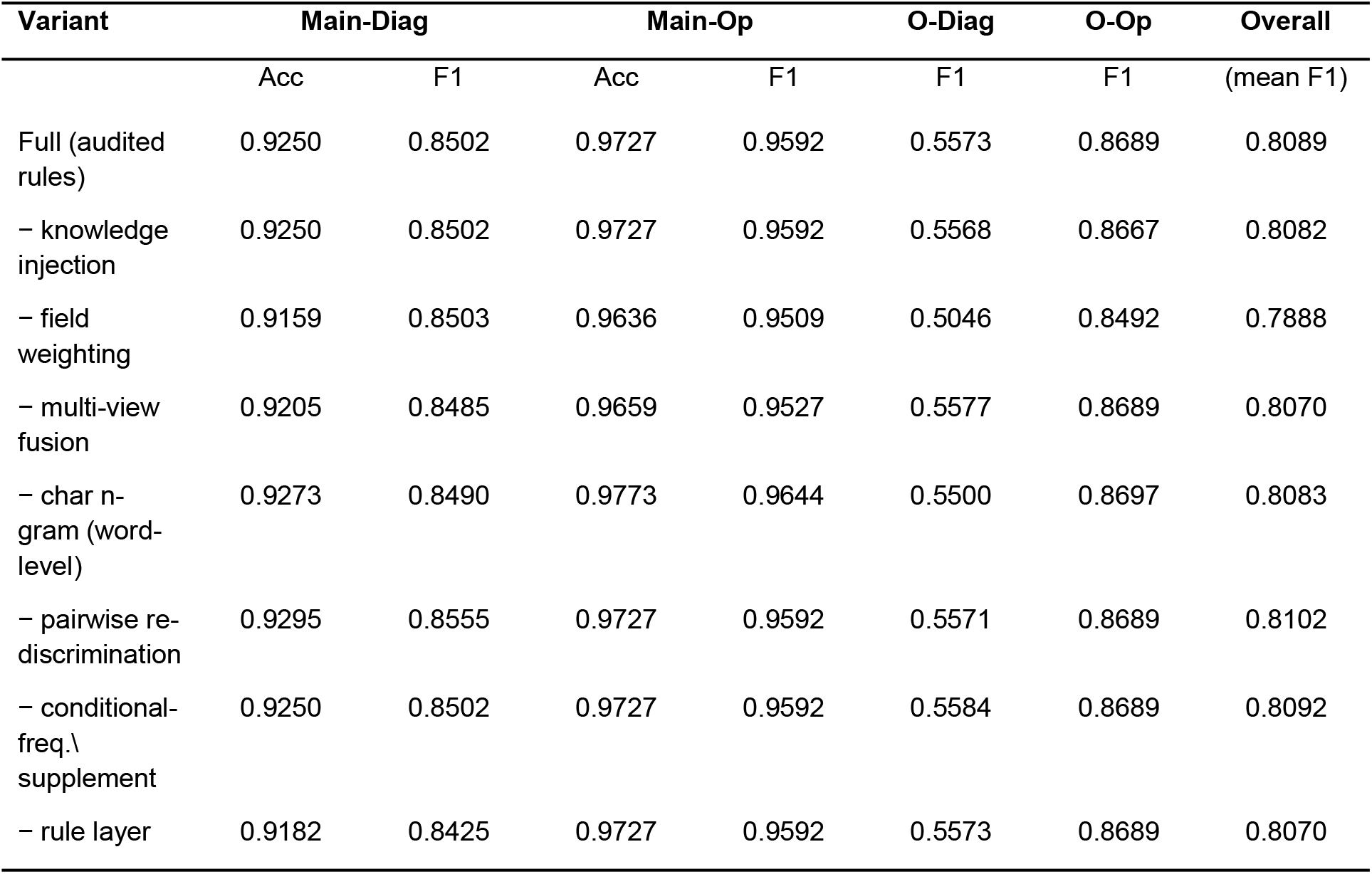
One-factor module ablations on the held-out split (8:2, seed 42), compared with the full framework. Removing field weighting is the largest single drop; knowledge injection, pairwise re-discrimination, conditional-frequency supplementation, and the character-vs-word choice are all within noise on this dataset.

| Variant | Main-Diag |  | Main-Op |  | O-Diag | O-Op | Overall<br>(mean F1) |
| --- | --- | --- | --- | --- | --- | --- | --- |
|  | Acc | F1 | Acc | F1 | F1 | F1 |  |
| Full (audited rules) | 0.9250 | 0.8502 | 0.9727 | 0.9592 | 0.5573 | 0.8689 | 0.8089 |
| – knowledge injection | 0.9250 | 0.8502 | 0.9727 | 0.9592 | 0.5568 | 0.8667 | 0.8082 |
| – field weighting | 0.9159 | 0.8503 | 0.9636 | 0.9509 | 0.5046 | 0.8492 | 0.7888 |
| – multi-view fusion | 0.9205 | 0.8485 | 0.9659 | 0.9527 | 0.5577 | 0.8689 | 0.8070 |
| – char n-gram (word-level) | 0.9273 | 0.8490 | 0.9773 | 0.9644 | 0.5500 | 0.8697 | 0.8083 |
| – pairwise re-discrimination | 0.9295 | 0.8555 | 0.9727 | 0.9592 | 0.5571 | 0.8689 | 0.8102 |
| – conditional-freq.\ supplement | 0.9250 | 0.8502 | 0.9727 | 0.9592 | 0.5584 | 0.8689 | 0.8092 |
| – rule layer | 0.9182 | 0.8425 | 0.9727 | 0.9592 | 0.5573 | 0.8689 | 0.8070 |

### 4.3 Comparisons: classical baselines and the deep reference point

Table 5 compares the frozen framework against classical baselines on the same splits and metrics, and against a fine-tuned Chinese pretrained encoder (MacBERT-base, 3 epochs, one seed). All baselines except MacBERT use plain single-view text (no field weighting, no injection). MacBERT reads key fields first (cut at 512 tokens), and its multi-label thresholds are tuned on the same validation split.

**Table 5.**
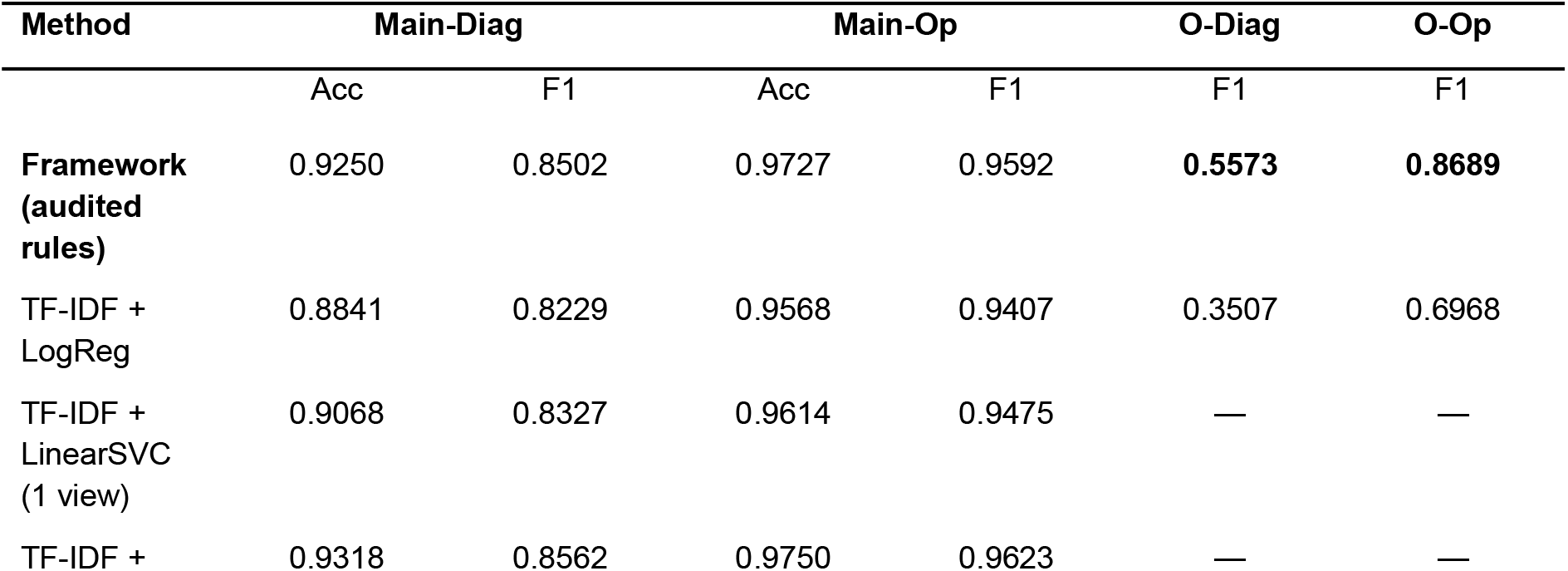
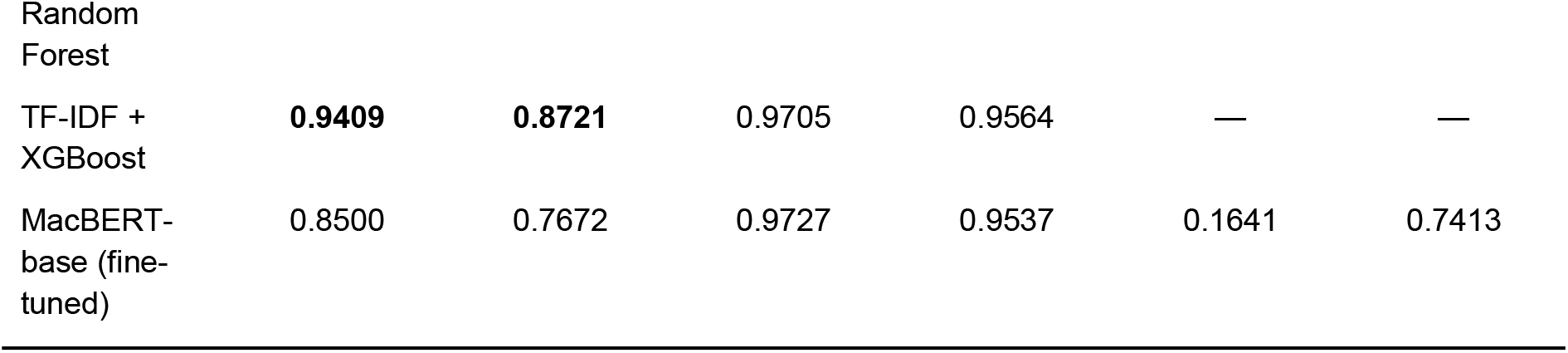
Baseline comparison on the held-out split (8:2, seed 42). Single-label tasks report Acc / macro-F1; multi-label tasks report sample-F1. The framework row is repeated from Table 1 for reference. RF/XGBoost were run only on the single-label tasks (cost in text); MacBERT thresholds were tuned on validation.

Compared with the single-view linear family, our design is worth +1.8/+1.1 points of single-label accuracy and +20.7/+17.2 points of multi-label sample-F1. The gain is largest on the multi-label tasks, but it is present on the head tasks too. Tree ensembles are competitive on the small head tasks: XGBoost gives the best main-diagnosis numbers (0.9409/0.8721) and Random Forest the best main-operation accuracy (0.9750). Their training time per task ranges from about 10–12 seconds (RF) to about 20 minutes (XGB) on CPU. However, they do not scale to the multi-label tasks under our cost budget. So the framework remains the best balanced system: it covers all four tasks in one ≈8-minute CPU run, and it is the only fully interpretable one. We report the head-task edge of tree ensembles plainly instead of hiding it. Most importantly, the deep reference point *falls behind at this data size*: fine-tuned MacBERT scores lower than the framework on all four tasks (main-diagnosis accuracy ™7.5 points; other-diagnosis sample-F1 0.164 vs.\ 0.557; other-operation 0.741 vs.\ 0.869), and ties only on main-operation accuracy. Yet it uses ≈505–510 s of GPU time and ≈6.7–6.9 GB of peak memory per task. The caveat is the single seed; multiple-seed repeats are left for follow-up work.

### 4.4 Data efficiency

Figure 2 shows the main-diagnosis learning curve (dual-view setting with codebook injection; 3 seeds per size). Accuracy rises from 0.876±0.041 at 550 records to 0.915±0.028 at 1,100 and 0.928±0.004 at 1,650. The full-protocol point is 0.923 (2,200 records, fixed paper split). Head-code performance therefore *saturates well before the full budget*: most of the reachable accuracy is already there at 1,100–1,650 records. This supports our data-access claim directly: a hospital with one to two thousand old records can expect near-plateau head-code performance. The large seed variance at small n (±3–6 points) shows that *which* records you have matters as much as how many at this scale. One methodological caveat: the sub-2,200 points re-train on random subsets with their own 20% evaluation splits, and macro-F1 is sensitive to the rare-class mix of small evaluation sets (this is why the nominal 0.894 at 1,650 is higher than 0.848 on the fixed 2,200 split). We therefore read the accuracy curve as the robust signal.

**Figure 2.**
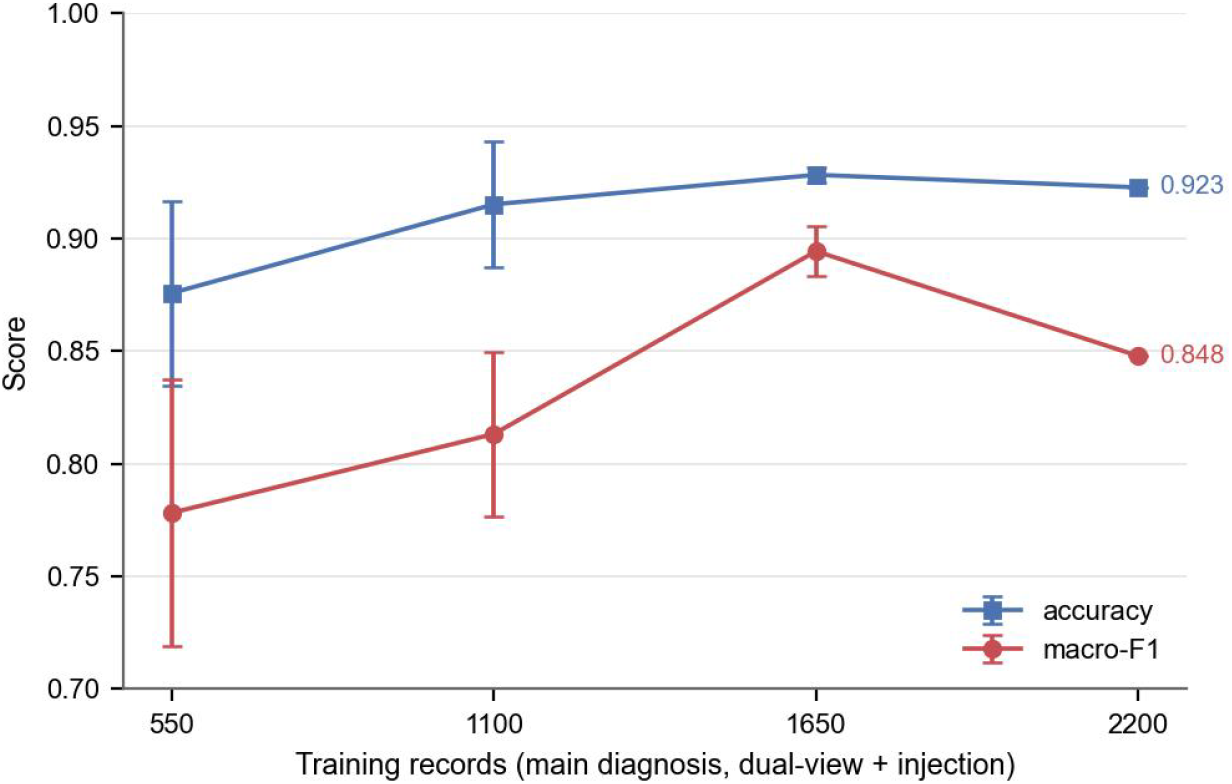
Main-diagnosis learning curve (mean ± std over 3 seeds; injection enabled). Accuracy saturates by roughly 1,650 records; the 2,200 point uses the fixed paper split.

### 4.5 Boundary, errors, efficiency, and interpretability

#### The honest boundary

Table 6 is the paper’s key honesty result. Head codes are solved at a usable level (other-operation F1 0.898). Mid-frequency codes drop sharply. Codes seen 1–2 times (about 61% of the other-diagnosis code space) are never predicted correctly. With at most two positive examples, a discriminative model cannot form a reliable boundary, and no threshold tuning fixes this. We therefore position the framework as *automation of frequent codes plus mandatory human review of rare codes*. We also argue that stratified reporting like Table 6 should be standard in low-resource coding studies, because headline F1 hides this failure mode.

**Table 6.**
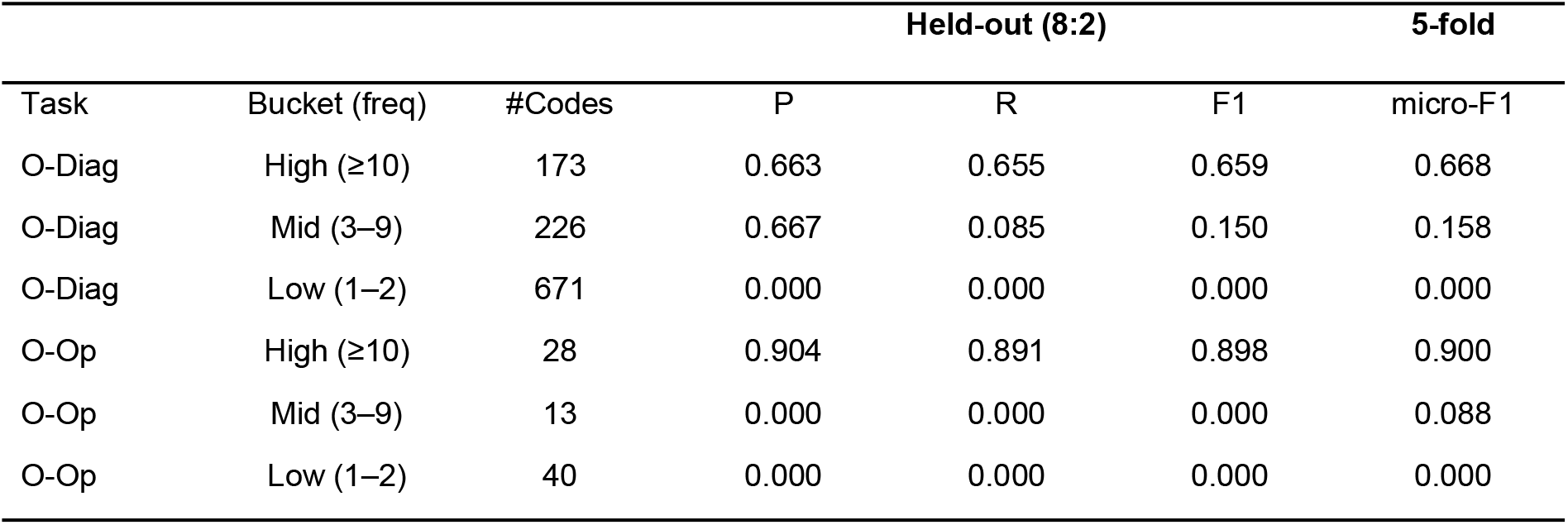
Frequency-stratified results for the multi-label tasks (buckets by training-set frequency of each code). Held-out split: micro-P/R/F1 per bucket; the 5-fold column reports micro-F1.

#### Errors

Main-diagnosis errors cluster in clinically adjacent code pairs: I63.900→I66.901 (5 cases), S32.000×002↔M80.900 (5+4), and Z51.100↔Z51.102 (4+4). The pairwise discriminator built for such pairs does not measurably reduce these errors; we flag confusion resolution as the clearest open weakness. The remaining main-operation errors are endoscopy-family distinctions (45.1300×004 vs.\ 45.1600×001). Other-diagnosis predictions under-predict: 3.44 codes per record vs.\ 4.31 gold (2.04 missing, 1.16 spurious). So the threshold trade-off still favors precision. Raising mid-frequency recall is the most valuable next step.

#### Efficiency

One 8:2 train-and-evaluate pass takes **489 s (≈8 minutes) on an ordinary desktop CPU**; a full 5-fold run takes 40–95 minutes depending on machine load. All six models together occupy 1.35 GB of RAM. The serialized files total 1.02 GB uncompressed and **493 MB with sparse-aware compression**. Per-record inference takes 13.7 ms (main-diagnosis view) and 24.0 ms (other-diagnosis model), well within interactive use. At this scale, the cost picture favors the framework: the deep baseline needs about half an hour of GPU time plus a multi-GB model download for the same four tasks.

#### Interpretability

A code’s decision score adds up from character n-gram contributions computed directly from the fitted coefficients, with no post-hoc surrogate. We observe three things. (i) Per-class evidence is the clinical wording itself: for K31.703 the top features are 息肉 (1.15), 胃息肉 (1.09), 胃底腺息肉 (0.96); for I63.900 they are 脑梗死 and 急性脑梗. (ii) For a rule-corrected case, the explanation of the pre-rule prediction Z51.102 is dominated by 肿瘤术后 n-grams. The rule and the explanation point to the same ambiguity, exactly what an auditable pipeline should expose. (iii) Injected anchors rarely dominate: the `DIAG_`c token of a correctly predicted class does not enter even the top-50 contributions. This mirrors the ablation null for codebook injection.

## 5. Advantages, Limitations, and Conclusion

### Advantages

(1) *Data*: the method trains on the few thousand records a hospital already owns, and head-code performance is near-plateau by 1,100–1,650 records. (2) *Compute*: the full four-task framework trains and evaluates in ≈8 minutes on an ordinary CPU, with no GPU and no pretrained weights, and predicts in 14–24 ms per record. (3) *Transparency*: every decision is deterministic and can be broken down into clinically readable n-gram evidence. (4) *Trust*: expert rules enter only through an audited channel with a measured fix/break record. We showed that naive clinical rules do more harm than good, and that pruning restores a safety check with no negative effect. (5) *Competitive at this scale*: with 1,760 training records, a fine-tuned Chinese pretrained encoder loses to the framework on all four tasks while needing a GPU, which reframes part of the low-resource gap as an algorithm problem.

### Limitations

(1) Long-tail failure: codes with 1–2 training examples stay at F1 = 0, so rare-code coding still needs human coders or few-shot / retrieval-augmented methods. (2) The evidence comes from one center and one language; cross-institution use requires re-validation on each hospital’s own data, and an external validation study is planned. (3) The deep comparison uses one seed and a base-size encoder on 1,760 records; larger encoders, longer training, and multi-seed repeats remain to be tested, and the tree baselines were run only on the single-label tasks. (4) Auditing rules on a single split risks overfitting that split; nested or repeated splits are the principled fix. (5) Field weights, confusable code pairs, and the rule list encode local clinical practice and may need re-tuning elsewhere; the audit method itself, however, is not tied to one institution.

## Conclusion

A carefully built classical pipeline (codebook injection, field-weighted multi-view construction, character n-gram linear models, and audited clinical rules) makes ICD coding workable under a real low-resource budget: usable head-code accuracy on a CPU, fully interpretable decisions, and expert rules that are safe because they are checked before use. We release the method and core code to support reproduction and local deployment in hospitals with limited resources.

## Data Availability

The data come from CCL2026-Eval Task 6 [9], constructed by the QLU-NLP group (Qilu University of Technology) and the First Affiliated Hospital of Shandong First Medical University. The data are governed by the competition’s data-use and confidentiality agreement: they cannot be redistributed by the authors, and they may be requested from the data providers under the evaluation’s terms. Core algorithm code is available from the authors within the scope of that agreement.

## Ethics Statement

This study is a retrospective secondary analysis of de-identified electronic medical records. The records were provided under the competition’s data-use and confidentiality agreement, which limits use to academic research and prohibits redistribution; the authors comply with all of its terms, including patient-privacy protection. The work contains no patient-identifiable information, no data records are redistributed, and the analysis involved no participant contact or intervention.

